# Sex-and-gender differences in cardiovascular risk factors and their correlates among adults in Freetown, Sierra Leone: A population-based health-screening survey

**DOI:** 10.1101/2023.03.15.23287178

**Authors:** James Baligeh Walter Russell, Theresa Ruba Koroma, Santigie Sesay, Sallieu K Samura, Sulaiman Lakoh, Ansu Bockarie, Onomeh Thomas Abiri, Joseph Sam Kanu, Lambert Tetteh Appiah, Joshua Coker, Abdul Jalloh, Victor Conteh, Sorie Conteh, Mohamed Smith, Durodami. R. Lisk

## Abstract

**Objectives:** to evaluate the association of sex-and-gender-specific cardiovascular disease risk factors, their prevalence, and correlates among adults in Sierra Leone.

**Study design:** This community-based cross-sectional study used a stratified multistage random sampling.

**Methods:** the survey was conducted in eight (8) selected randomized sub-zonal communities across the western urban area in Sierra Leone, with an included sampling of 2394 adults. The WHO stepwise approach for non-communicable diseases was utilized. Multivariable logistic regression was done to determine associations between demographic characteristics and cardiovascular risk factors.

**Results:** The prevalence of hypertension (33.4% vs 37.4%, p=0.068), diabetes mellitus (7.4% vs 9.2%, p=0.101), overweight (32.3% vs 34.2%, p=0.323) and obesity (9.9% vs 10.2%, p=0.818) were higher among males in comparison to females. Body Mass Index (BMI) (25.0 ± 5.0 vs 24.6 ± 4.4, p = 0.029), waist circumference (WC) (93.6 ± 4.5 vs 80.0 ± 5.0, p < 0.001), triglyceride (1.7±0.35 vs 1.6±0.32, p=0.013), total cholesterol (5.1±0.77 vs 4.9±0.66, p < 0.001) and low HDL-C (1.28 ± 0.29 vs 1.3±0.24, p = 0.016) were significantly higher among females as compared to males. The odds of having dyslipidemia [OR = 1.339; 95% C.I: (1.101-1.629), p=0.003] and consuming alcohol [OR = 1.229; 95% C.I: (1.026-1.472), *p*=0.025] were higher among females. Women had 1.8 times greater odds [AOR=1.849; 95% C.I: (0.713 - 1.010), *p*=0.030] of being hypertensive, 1.4 times greater odds [AOR=1.441; 95% C.I: (1.176 - 1.765), *p*=<0.001] of being dyslipidemic and 1.2 times greater odds [AOR = 1.225, 95% C.I: (1.0123-1.481), *p*=0.037] of consuming alcohol compared to men. BMI, WC, and raised blood sugar had a strong correlation among women than men.

**Conclusion:** Being female was associated with a high prevalence of cardiovascular health risks in Sierra Leone. This study emphasizes the importance of reducing the CVD burden among females through policies related to public health education and screening strategies.

## 1. Background

Over the past decades, public health research in cardiovascular medicine was primarily focused on men, with “sex and gender” differences in the risk factors of cardiovascular diseases being grossly underestimated. [1,2]. Recently, enormous efforts through scientific researchers and Women’s Health Initiatives have improved our understanding not only of the concept of “sex and gender” differences in cardiovascular disease CVD but also of the recognition of heart disease in women [3,4,5]. Hence, the “one size fits all for men approach’ may not hold.

Sex is a “biological variable” that is genetically determined. In contrast, gender is a “multifactorial social concept” that is linked to customs, religions, traditions, public beliefs, and individual behaviours that exists within a historical and social setting” [6]. The differences in sex hormones and chromosomes between males and females are the biological factors that may protect a female against hypertension until menopause, but thereafter, gender differences in hypertension remained negligible [7]. Behavioural factors contributing to the difference between males and females include body mass index (BMI), smoking, alcohol consumption, unhealthy diet intake and low physical activities. While smoking is more common in men, it is reported to have more deleterious consequences on females than males because of gender differences in nicotine metabolism. However, the gender difference in smoking has significantly narrowed in recent decades. [8]. Studies have shown that the risk of developing ischemic heart disease or stroke is 25% higher among female smokers than male smokers [9,10]. Unlike men, women are more likely to eat healthy diets but are less physically active than men. [5].

In Africa, there is a dearth of data on “sex and gender-related” differences for most cardiovascular risk factors. [11,12]. Despite the increasing burden of cardiovascular disease in sub-Saharan Africa (SSA), the relationship between sex/gender and CVD is unknown. In this research area, studies from West Africa are limited, and there are no published studies from Sierra Leone. We, therefore, aimed to assess sex-and-gender differences in the prevalence of cardiovascular risk factors and their correlates among adults in Freetown, Sierra Leone, using identifiable cardiovascular risk factors from the Ecobank (Sierra Leone) study database. Our findings will provide information to assist future research, potential prevention, and therapeutic intervention against non-communicable diseases (NCDs).

## 2. METHODS

### Ethical approval and registration

The Sierra Leone Ethics and Scientific Review Committee approved the research protocol, questionnaire, and consent form. The protocol of this study was registered under Research Registry with the unique identification number researchregistry8201 and is available at https://www.researchregistry.com/browse-the-registry#home/. Serial-coded numbers were assigned to the case records to maintain anonymity, and all data extracted was handled with strict confidentiality. This study was written according to the STROBE (Strengthening the Reporting of Observational Studies in Epidemiology) statement guidelines [13].

### 2.1. Study design, setting and cohort group

A population-based cross-sectional study was conducted between October 2019 and October 2021 among adults in West Area Urban, Freetown, Sierra Leone. Ecobank Sierra Leone Limited funded this screening and awareness programme for NCD. Freetown, Sierra Leone’s capital, has a dense heterogenous population of an estimated 1.5 million inhabitants [22]. It was selected as the study site because it serves as the primary business centre in the country, and its demographic distribution is represented by all ethnic groups, with Krio and English being the primary spoken language.

### 2.2. Sample size, recruitment, and selection

This study used the “Ecobank health screening and awareness programme” data for NCD in Freetown, Sierra Leone. A stratified random sampling strategy was used to recruit adult Sierra Leonean participants aged ≥ 20. The first stage in the sampling was to select all official electoral constituencies (Central I, II, East I, II, III, & West I, II, III) in Western Area - Urban, and then followed by subdividing each constituent region into subzones using the 2015 census data [14]. Upon selecting a sub-zonal community from an official electorate by simple random sampling, the potential line-listed participants were further selected at their community health centre by simple random sampling methods. Pregnant/lactating mothers, participants with mental illness/dementia and persons unwilling to grant consent were excluded from the study. The sample size of this study was calculated by the Leslie Kich formula, using the clinical estimated prevalence of 22% for hypertension in Sierra Leone, with a 5% precision and 95% Confidence Interval [15, 16]. By minimizing bias and allowing attrition of non-response and non-availability of data, the study was oversampled by 20%. Using a design effect of the eight sub-zonal communities, the adjusted estimated sample size was 2531.

Eligible participants from each selected sub-zonal community were invited to the National Victoria park in Freetown for participation in the “awareness and screening campaign for non-communicable disease”. Medical students, doctors and nurses were trained on the campaign’s conduct, including data collection. At the health screening venue (Victoria Park), consented and enrolled participants who had completed their screening questionnaires and anthropometric measurements were referred to an accredited reference laboratory for blood sample collection. (Flow chart during the campaign is shown in Fig:1)

### 2.3. Data Collection and Measurements

#### Demographic and Behavioural Characteristic Data

Three categories of data collection were done using the WHO stepwise approach for NCD. Socio-demographic and behavioural characteristics information were collected using a pre-tested face-to-face interview questionnaire. Socio-demographics collected were age, sex, gender, educational level, religion, marital status, and medical history (family history of hypertension and diabetes mellitus). At the same time, behavioural characteristics, including fruit and vegetable consumption, smoking status, alcohol consumption and physical activity, were collected under supervision. Physical activity was assessed and classified into “Low”, “Moderate”, and “Vigorous” by using the WHO physical activity form [17]. Data on alcohol consumption was based on the WHO step survey tool [18]. A participant who smoked more than 100 sticks of cigarettes in their lifetime and, or still smoking at the time of the interview, was referred a smoker, while an ex-smoker is someone who had stopped smoking at least 28 days before the interview [19]. Translators assisted participants who could not understand English during data collection.

### 2.4. Anthropometric measurements and biochemical analyses

The systolic blood pressure (SBP) and diastolic blood pressure (DBP) of each participant were recorded on the right arm while in the sitting position by using an electronic blood pressure monitor (OMRON M3). After at least 3-5 minutes of rest, two consecutive measurements were recorded, and the mean was taken as the final BP reading. Hypertension was defined as an average SBP of 140 mm Hg or higher, or DBP of 90 mm Hg or greater or a participant reporting current use of antihypertensive medication [20]. Body weight, height, and waist circumference were measured with light clothes and bare feet. Weight was measured to the nearest 0.1 kilograms, while Height was measured by a stadiometer closest to 0.01 meter. The body mass index (BMI) was calculated as a ratio of the weight in kilograms and the square of the height in metres. BMI-based body habitus (in kg/m^2^) was classified as underweight (BMI <18.5), normal weight (BMI=18.5–24.9), overweight (BMI=25.0–29.9) and obese (BMI ≥30) [21].

A tape measure recorded waist circumference (WC) at the approximate centre between the lower margin of the last palpable rib and the top of the iliac crest. Abdominal obesity was defined as a waist circumference > 88 cm for women and 102 cm for men [22].

### 2.5. Clinical biochemistry measurements

Participants’ blood samples were collected from the median cubital vein between 8:00 and 10:00 AM after overnight fasting for 8 to 10 hours. These samples were processed within 4 hours of collection per manufacturers’ instructional protocols, using Beckman Coulter: AU480 Chemistry System. Glucose, total cholesterol, triglycerides, high-density lipoprotein (HDL-C) and low-density lipoprotein (LDL-C) were analysed.

Diabetes mellitus was defined as a fasting blood glucose (FBG) level of 7.0 mmol/L or greater, HbA1c ≥ 6.5%, or using insulin or an oral hypoglycaemic agent. Pre-diabetes was defined as FPG between 6.1mmol/l (110 mg/ dL) and 6.9 mmol/l (124.9 mg/dL) [23].

Dyslipidaemia was defined as TG ≥ 1.70 mmol/L (150 mg/dL), TC ≥ 6.22 mmol/L (240 mg/dL), LDL ≥ 3.3mmol/L (130 mg/dL), HDL <1.04 mmol/L (40 mg/dL) or the use of lipid-lowering medications, was considered as abnormally high. [24].

### 2.6. Statistical Analysis

Data analysis was done using IBM SPSS Statistical 2.6 and STATA 17 software. Baseline characteristics, cardiometabolic risk factors and target organ damage characteristics were analyzed by sex and zones. Categorical variables were expressed as numbers and percentages, and the Pearson chi-square (X^2^) test was used to assess the difference. Continuous variables were expressed as mean ± SD and compared using a one-way analysis of variance (ANOVA). Median and IQR were used when necessary. Multivariable logistic regression was done to determine associations between demographic characteristics and cardiovascular risk factors. A two-tailed p-value of ≤ 0.05 was considered statistically significant.

## 3. RESULTS

Of the 2531 participants that met the eligibility criteria, only 2394 individuals were recruited into the study, and the response rate was 94.6%. Fifty-four individuals could not attend the “Ecobank - cardiovascular health screening campaign” day at the National Victoria Park, while 76 participants could not complete laboratory and cardiovascular screening. There was an equal representation from the eight sub-zonal communities without significant differences in population distribution (p=0.950).

### 3.1. Socio-demographic and behavioral profile of the study sample

Descriptive socio-demographic characteristics of participants for the population are summarized in Table 1. The mean age of the study participants was 41.9 ± 12.3 years with an age range of 20 to 75 years. The mean age of females (42.06 ± 12 years) was not statistically different from that of males (41.76 ± 12.6 years). The results showed low earning income (0 - 500 Leones) in the majority (38.4%) of the participants, with no significant differences between women and men (54.8% of women vs 41.6% of men, p = 0.052). More females, compared to men, attained primary and secondary education, but more men were likely to graduate from tertiary institutions. These differences between men and women in achieving education were insignificant (p = 0.095). Most of the study participants were unemployed (38.8%), with a statistical difference among self-employed female participants. [men (45.4%) vs (54.6%), p < 0.001]. Even though the study suggested women were significantly more likely to consume alcohol (p = 0.014), daily alcohol intake was significant among men (p = 0.046).

**Table 1:** Gender differences in socio-demographic characteristics of participants.

| Characteristics | Total n(%) | Female n(%) | Male n(%) | P-value($\chi^2$) |
| --- | --- | --- | --- | --- |
| Basic characteristics |  |  |  |  |
| No. | 2394 | 1250 | 1144 |  |
| Age, by group |  |  |  |  |
| 20-29 | 447(18.7) | 255(57.0) | 192(43.0) | <0.001(28.9) |
| 30-39 | 703(29.4) | 383(54.5) | 320(45.5) |  |
| 40-49 | 684(28.6) | 359(52.5) | 325(47.5) |  |
| 50-59 | 356(14.9) | 141(39.6) | 215(60.4) |  |
| >60 | 204(8.5) | 112(54.9) | 92(45.1) |  |
| INCOME (Currency = Leone) |  |  |  |  |
| 0-500 | 920(38.4) | 504(54.8) | 416(45.2) | 0.052(7.715) |
| 500-1000 | 666(27.8) | 319(47.9) | 347(52.1) |  |
| 1100-2000 | 490(20.5) | 256(52.2) | 234(46.2) |  |
| >2000 | 318(13.3) | 171(53.8) | 147(46.2) |  |
| Education level |  |  |  |  |
| None | 618(25.8) | 318(51.5) | 300(48.5) | 0.095(6.361) |
| Primary | 479(20.0) | 273(57.0) | 206(43.0) |  |
| Secondary | 835(34.9) | 432(51.7) | 403(48.3) |  |
| Tertiary | 462(19.3) | 227(49.1) | 235(50.9) |  |
| Marital status |  |  |  |  |
| single | 955(39.9) | 504(52.8) | 451(47.2) | 0.699(1.428) |
| Married | 737(30.8) | 372(50.5) | 365(49.5) |  |
| Separated/Divorce | 586(24.5) | 310(52.9) | 276(47.1) |  |
| Widow | 115(4.8) | 63(54.8) | 52(45.2) |  |
| Occupation |  |  |  |  |
| Employed | 500(20.9) | 302(60.4) | 198(39.6) | <0.001(27.6) |
| Self-employed | 531(22.2) | 241(45.4) | 290(54.6) |  |
| Unemployed | 930(38.8) | 466(50.1) | 464(49.9) |  |
| Retired | 167(7.0) | 96(57.5) | 71(42.5) |  |
| Student | 264(11.0) | 145(54.9) | 119(45.1) |  |
| BLOOD PRESSURE (mmHg) |  |  |  |  |
| Normal | 1061(44.3) | 510(48.1) | 551(51.9) | 0.002(15.15) |
| Pre-hypertension | 489(20.4) | 278(56.9) | 211(43.1) |  |
| Hypertension stage 1 | 644(26.9) | 360(55.9) | 284(44.1) |  |
| Hypertension stage 2 | 200(8.4) | 102(51.0) | 98(49.0) |  |
| FRUITS/VEGETABLE |  |  |  |  |
| <3 serving | 2182(91.1) | 1143(52.4) | 1039(47.6) | 0.592(0.28) |
| >3 serving | 212(8.9) | 107(50.5) | 105(49.5) |  |
| Alcohol |  |  |  |  |
| Never | 1486(62.1) | 783(52.7) | 703(47.3) | 0.014(8.14) |
| Current | 652(27.2) | 316(48.5) | 336(51.5) |  |
| Previous | 256(10.7) | 151(59.0) | 105(41.0) |  |
| Alcohol intake |  |  |  |  |
| Daily | 345(14.4) | 151(43.8) | 194(56.2) | 0.046(6.14) |
| Weekly | 285(11.9) | 150(52.6) | 135(47.4) |  |
| Monthly | 134(5.6) | 57(42.5) | 77(57.5) |  |
| Smoking |  |  |  |  |
| Never | 2073(86.6) | 1088(52.5) | 985(47.5) | 0.797(0.45) |
| Current | 198(8.3) | 100(50.5) | 98(49.5) |  |
| Ex-smoker | 123(5.1) | 62(50.4) | 61(49.6) |  |
| Daily physical activity |  |  |  |  |
| Low | 895(37.4) | 481(53.7) | 414(46.3) | 0.002(12.38) |
| Moderate | 939(39.2) | 515(54.8) | 424(45.2) |  |
| Vigorous | 511(21.3) | 233(45.6) | 278(54.4) |  |

Table 1 Continued
| Characteristics | Total n(%) | Female n(%) | Male n(%) | P-value($\chi^2$) |
| --- | --- | --- | --- | --- |
| Measures of adiposity |  |  |  |  |
| BMI (kg/m<sup>2</sup>) |  |  |  |  |
| Underweight | 38(1.6) | 21(55.3) | 17(44.7) | 0.066(7.2) |
| Normal | 1502(62.7) | 794(52.9) | 708(47.1) |  |
| Overweight | 612(25.6) | 329(53.8) | 283(46.2) |  |
| Obese | 240(10.0) | 106(44.2) | 134(55.8) |  |
| Waist circumference ($\geq 94$cm men, $\geq 80$cm women) | | | | |
| Normal | 1882(78.6) | 870(46.2) | 1012(53.8) | 0.003(8.569) |
| Abnormal | 512(21.4) | 274(53.5) | 238(46.5) |  |
| WHtR risk |  |  |  |  |
| Normal ($\leq 0.5$) | 1050(43.9) | 36(3.4) | 1014(96.6) | <0.001(1786.73) |
| Increased risk (0.51- 0.59) | 1276(53.3) | 1146(89.8) | 130(10.2) |  |
| High risk ($>0.6$) | 68(2.8) | 68(100) | 0(0.0) | |
| WHR ($\geq 0.90$ men, $\geq 0.85$ women) | | | | |
| Low | 763(31.9) | 753(98.7) | 10(1.3) | 0.210(3.12) |
| Moderate | 273(11.4) | 271(99.3) | 2(0.7) |  |
| High | 207(8.6) | 207(100) | 0(0.0) |  |
| Laboratory |  |  |  |  |
| Lipids |  |  |  |  |
| Total Cholesterol (TC) ($\geq 6.2$mmol/l) | | | | |
| Normal | 2163(90.4) | 1171(54.1) | 992(45.9) | <0.001(32.25) |
| High | 231(9.6) | 79(34.2) | 152(65.8) |  |
| LDL-C ($\geq 3.3$mmol/l) | | | | |
| Normal | 2077(86.8) | 1083(52.1) | 994(47.9) | 0.858(0.03) |
| High | 317(13.2) | 167(52.7) | 150(47.3) |  |
| HDL-C ($\leq 1.04$mmol/l) | | | | |
| Normal | 2129(88.9) | 1142(53.6) | 987(46.4) | <0.00115.68) |
| High | 317(13.2) | 108(40.8) | 157(59.2) |  |
| TRIGLYCERIDE ($\geq 1.7$mmol/l) | | | | |
| Normal | 1862(77.8) | 1011(54.3) | 851(45.7) | <0.001(14.58) |
| High | 530(22.1) | 238(44.9) | 292(55.1) |  |
| DIABETES (mmol/l) |  |  |  |  |
| Normal ($>6$mmol/l) | 2084(87.1) | 1080(51.8) | 1004(48.2) | 0.234(2.9) |
| Pre-diabetes (6.0 – 6.9mmol/l) | 103(4.3) | 51(49.5) | 52(50.5) |  |
| Diabetes ($>7.0$mmol/l) | 199(8.3) | 115(57.8) | 84(42.2) | |

Compared with their male counterparts, low daily physical activity was significantly higher in women (p = 0.002). Fruit intake of 8.9% was very low among the participants, without a significant difference among gender (p = 0.592). There was no gender difference in the smoking rate in the study population (p = 0.797).

### 3.2. Gender Differences in Risk Factors for Cardiovascular Diseases

Gender differences in cardiovascular risk factors in males and females are summarized in table 2. Women had significantly higher mean BMI, waist circumference, total cholesterol, triglyceride and low HDL-C. On the contrary, age, weight, height, systolic blood pressure, diastolic pressure, LDL-C, FBS and HBA1C were not significantly different by gender.

**Table 2:** Gender differences in cardiovascular risk factors in mean.

| Characteristics | Total (2394) | Female (1144) | Male (1250) | p-value |
| --- | --- | --- | --- | --- |
| | N±SD | Mean ± SD | Mean ± SD | |
| Age | 41.9±12.3 | 42.06±12.0 | 41.76±12.6 | 0.550 |
| Weight | 74.3±14.9 | 74.8±15.6 | 73.8±14.2 | 0.78 |
| Height | 1.7±0.05 | 1.7±0.05 | 1.7±0.05 | 0.302 |
| BMI | 24.8±4.7 | 25.0±5.0 | 24.6±4.4 | **0.029** |
| WC | 87.1±8.3 | 93.6±4.5 | 80.0±5.0 | **< 0.001** |
| SBP | 127.8±23.3 | 127.5±23.4 | 128.1±23.2 | 0.516 |
| DBP | 85.7±11.3 | 85.4±11.6 | 86.0±11.0 | 0.188 |
| Triglyceride | 1.65±0.34 | 1.7±0.35 | 1.6±0.32 | **0.013** |
| Total Cholesterol | 5.0±0.72 | 5.1±0.77 | 4.9±0.66 | < **0.001** |
| Low HDL-C | 1.3±0.27 | 1.28±0.29 | 1.3±0.24 | **0.016** |
| LDL-C | 3.0±0.66 | 2.98±0.66 | 3.0±0.65 | 0.315 |
| FBS | 5.1±1.6 | 5.0±1.61 | 5.2±1.66 | 0.080 |
| HBA1C | 5.2±1.03 | 5.2±0.96 | 5.2±1.09 | 0.964 |

The prevalence of cardiovascular risk factors by gender is represented in Table 3. The prevalence of hypertension and diabetes mellitus was 35.3% and 8.3%, respectively. Overweight (33.2%), obesity (10%), alcohol consumption (37.9%), abnormal waist circumference (21.4%) and lack of physical activity (37.4%) were common. Among women, the prevalence of higher waist circumference, elevated total cholesterol, high triglyceride, and low physical activities were significantly greater than in men.

**Table 3:** Prevalence of cardiovascular risk factors according to sex.

| Characteristics | Total n(%) | Female n(%) | Male n(%) | p-value |
| --- | --- | --- | --- | --- |
| Overweight | 796(33.2) | 369(32.3) | 427(34.2) | 0.323 |
| Obese | 240(10.0) | 113(9.9) | 127(10.2) | 0.818 |
| Abnormal waist circumference | 512(21.4) | 274(24.0) | 238(19.0) | **0.003** |
| Hypertension | 844(35.3) | 382(33.4) | 462(37.4) | 0.068 |
| Alcohol intake | 908(37.9) | 467(37.4) | 441(38.5) | 0.025 |
| Hx of smoking | 195(8.3) | 103(9.2) | 92(7.5) | 0.139 |
| Diabetes | 199(8.3) | 84(7.4) | 115(9.2) | 0.101 |
| High Cholesterol | 231(9.6) | 152(13.3) | 79(6.3) | <**0.001** |
| High LDL-C | 317(13.2) | 150(13.1) | 167(13.4) | 0.858 |
| High Triglyceride | 530(22.2) | 292(25.5) | 238(19.1) | <**0.001** |
| Low daily physical activity | 895(37.4) | 481(40.1) | 414(39.0) | **0.002** |

Using the univariate and multivariate logistic regression for gender and cardiovascular risk factors, the crude and adjusted odds ratios were summarized in Table 4. The odds of being overweight, obesity, smoking, having hypertension, diabetes, physical inactivity, and fruit/vegetable consumption were not statistically significant by gender. However, after adjusting for hypertension, dyslipidemia and alcohol, women had 1.8 times greater odds of being hypertensive, 1.4 times greater odds of being dyslipidemic and 1.2 times greater odds of consuming alcohol than men.

**Table 4:** Univariate and Multivariate logistic regression of gender and cardiovascular risk factors.

| Variables | Univariate OR |  | Multivariate OR |  |
| --- | --- | --- | --- | --- |
|  | COR 95% CI | p-value | AOR 95%CI | p-value |
| **Overweight** |  |  |  |  |
| No | Ref. |  | Ref. |  |
| Yes | 0.918(0.774 - 1.088) | 0.323 | 1.018(0.831 - 1.246 ) | 0.079 |
| **Obese** |  |  |  |  |
| No | Ref. |  | Ref. |  |
| Yes | 0.968(0.741 - 1.266) | 0.818 | 0.941(0.686 - 1.290) | 0.526 |
| **Smoking** |  |  |  |  |
| Never | Ref. |  | Ref |  |
| Yes | 1.248(0.930 - 1.673) | 0.802 | 1.269(0.940 - 1.712) | 0.158 |
| **Hypertension** |  |  |  |  |
| No | Ref. |  | Ref |  |
| Yes | 0.855(0.723 - 1.012) | 0.068 | 1.849(0.713 - 1.010) | **0.030** |
| **Dyslipidemia** |  |  |  |  |
| No | Ref. |  | Ref. |  |
| Yes | 1.339(1.101 - 1.629) | **0.003** | 1.441(1.176 - 1.765) | <**0.001** |
| **Diabetes Mellitus** |  |  |  |  |
| No | Ref. |  | Ref. |  |
| Yes | 0.782(0.583 - 1.049) | 0.101 | 0.826(0.611 - 1.117) | 0.135 |
| **ALCOHOL** |  |  |  |  |
| No | Ref. |  | Ref. |  |
| Yes | 1.229(1.026 - 1.472) | **0.025** | 1.225(1.0123 - 1.481) | **0.037** |
| **Physical activity** |  |  |  |  |
| No | Ref. |  | Ref. |  |
| Yes | 1.090(0.927 - 1.289) | 0.310 | 1.090(0.919 - 1.293) | 0.323 |
| **Fruits/Vegetable intake** |  |  |  |  |
| <3 servings | Ref. |  | Ref. |  |
| >3 servings | 1.079(0.814 - 1.431) | 0.595 | 1.063(0.795 - 1.421) | 0.682 |

### 3.3. Correlates of CVD Risk Factors

The correlates of CVD risk factors are represented in Figure 2. Among women, body mass index, waist circumference and FBS were significantly correlated with CVD risk factors.

**Figure 1:**
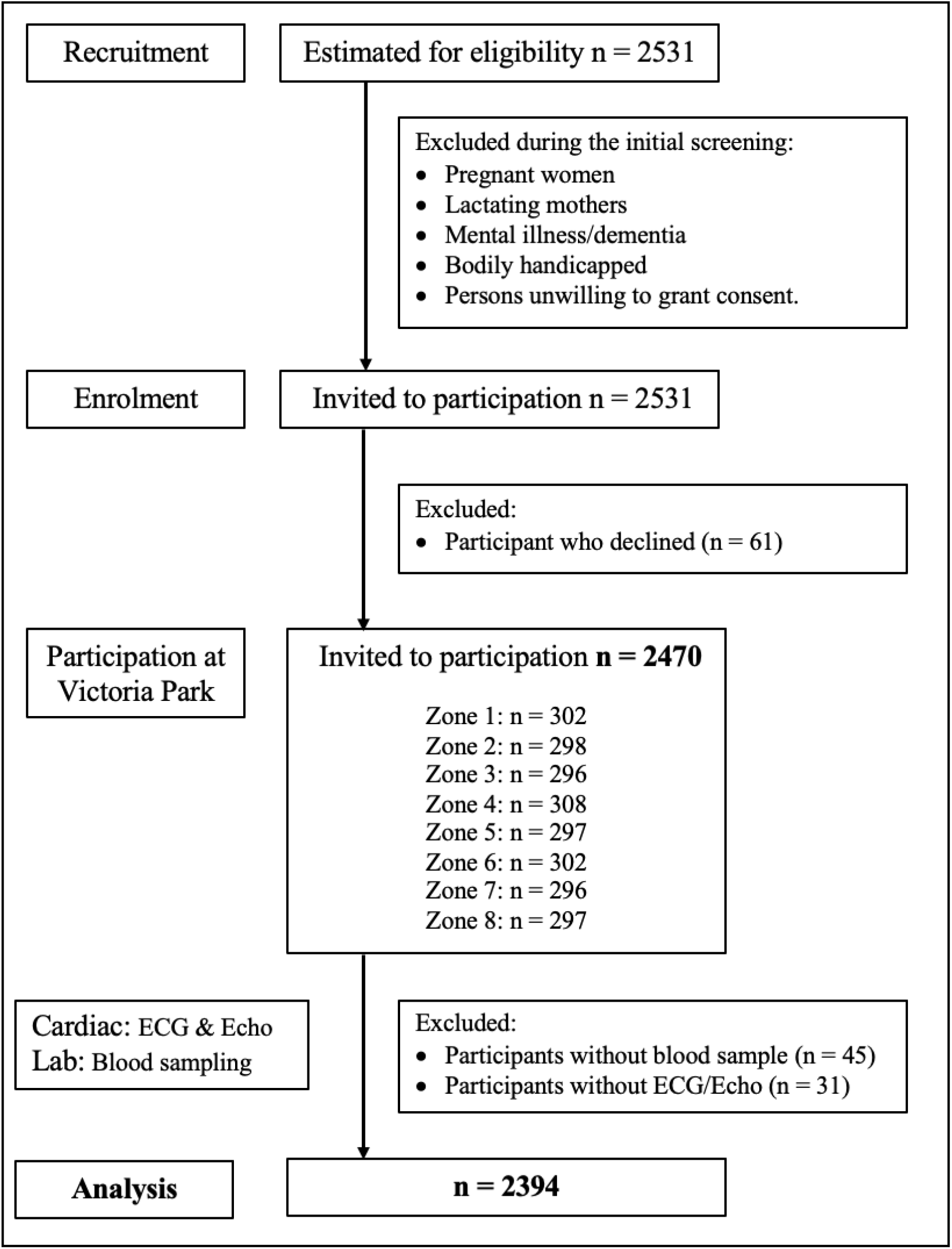
Steps involved during recruitment of participants and final analysis of data.

**Figure 2:**
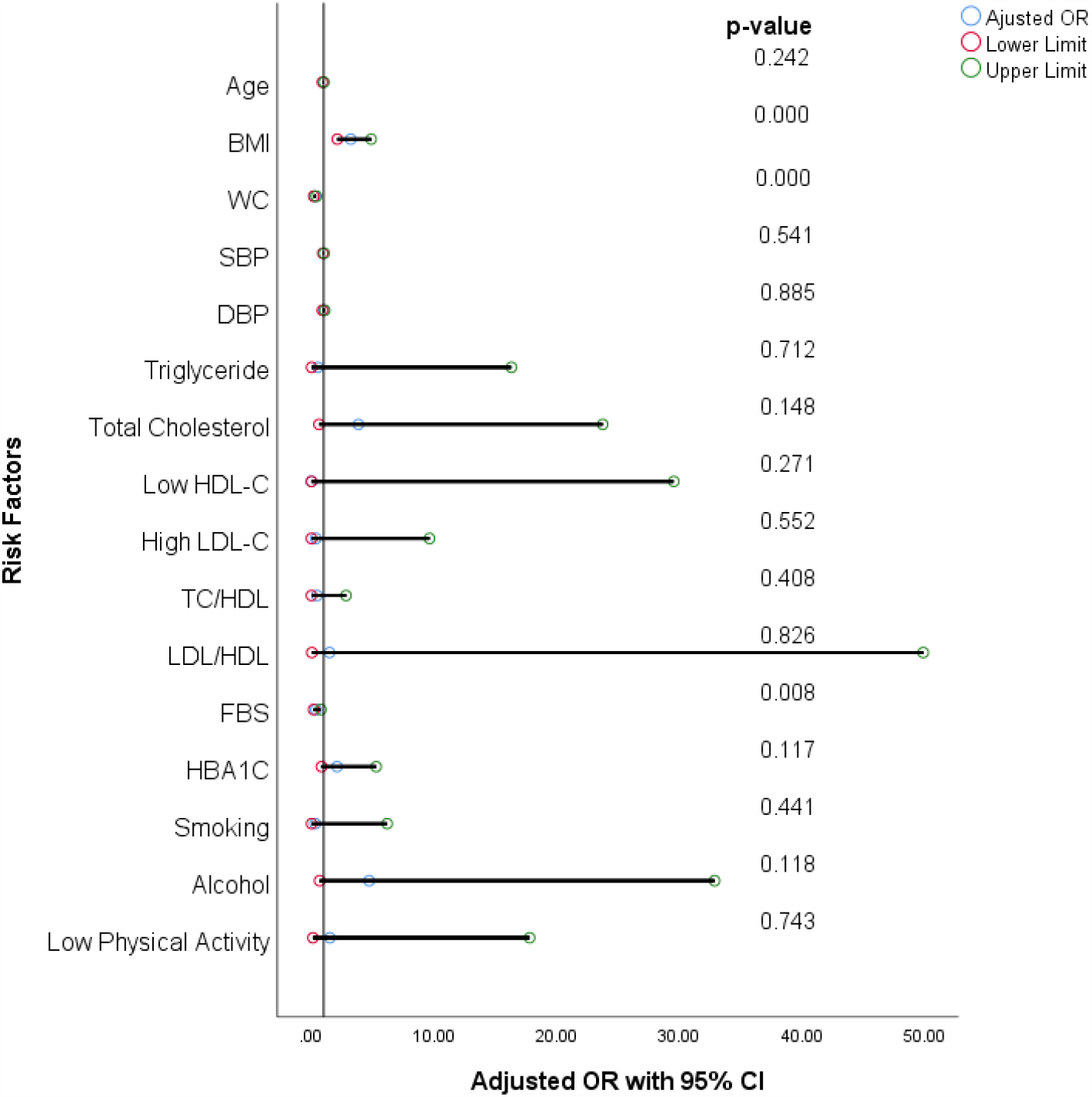
Correlates of cardiovascular risk factors among participants

## 4. Discussion

Sex and gender disparities in cardiovascular risk factors are relevant not only to developed countries but to low- and middle-income countries (LMIC) countries where the disease burden is increasing. There are limited scientific reports from SSA despite several health screening campaigns on cardiovascular diseases conducted in this region. This health screening and awareness campaign provided the largest representative data on sex and gender disparity for NCD among adults in Sierra Leone. This is the first report describing sex-gender differences in the prevalence of cardiovascular risk in an adult population in West Africa.

In this study, we reported a high prevalence of cardiovascular disease risk factors: hypertension (35.3%), diabetes mellitus (8.3%), Overweight (33.2%), Obesity (10.0%), abdominal obesity (21.4%), dyslipidaemia (21.4%), low physical inactivity (37.4%), smoking (8.3%) and alcohol consumption (37.9%). The study also documented gender disparity in several CVD risk factors, mainly increased BMI and abdominal obesity, as well as raised total cholesterol, triglyceride, and low HDL. The high prevalence of excess body weight among women in our study is comparable to other findings from Cameroon, Kenya, and Tanzania [11,25-27].

In Africa, it is widely accepted that the increasing number of overweight/obesity in women in an urban setting is attributed to socio-cultural influences, like the gender-specific pattern of work, sedentary lifestyle, and physical attractiveness of being fat among women. [28, 29]. Contrary to our study, a high prevalence of obesity among men compared to women has been reported in Mexico and China [30, 31].

The high prevalence of hypertension among men in our study is consistent with findings reported by Sandberg et al and Njelekela et al but in contrast with the results documented by Dev et al. [7,27,32]. The variation in gender disparity in hypertension among reported studies could be attributed to differences in the study designs, socio-demographic characters, and lifestyle patterns of the study participants. Since two-thirds of our study participants in the cohort were below the age of 50 years, the high prevalence of hypertension among men could be attributable to the biological factor (oestrogen) that protect female against hypertension until menopause. Even though BMI, abdominal obesity and WHtR were significantly raised among women, the risk of hypertension was lower among women in our study. Similar findings have been reported in the Tanzanian study by Njelekela et al. [27].

The association between hypertension and increased BMI is well established in urban settings in SSA and is partly associated with increased cardiac output and arterial resistance [33,34]. An increased body weight of 10kg may result in a 3.0 mmHg higher systolic blood pressure and a 2.3 mmHg higher diastolic blood pressure [35]. For our obese female participants in this study, it would be assumed that a greater degree of adiposity must be achieved to attain a significant rise in blood pressure. Nonetheless, the increased prevalence of obesity in this young female population is worrying as it is associated with an increased risk of cardiovascular risk. We observed significant gender-specific differences in cardiovascular disease risk factors among women for TC, triglyceride and low HDL. Our findings are consistent with reports from other African studies [36,37]. The gender disparity of this lipid profiling of CVD risk factors could be attributed to the increased adiposity documented among females.

The prevalence of type 2 diabetes mellitus is reported to be trending on the increase in Sierra Leone – 2.4% in 1997, 3.5% in 2009, 6.2% in 2017, and 5.5% in 2020 [46,47,48]. Our study has documented the highest prevalence (8.3%) of diabetes mellitus in Sierra Leone, with a high prevalence of diabetes mellitus in females (9.2%) than in males (7.5%). It is the first study in Sierra Leone to measure HBA1c in the screening for diabetes, and its combination with FBS could have accounted for the high prevalence of diabetes mellitus among adults in an urban setting.

There are genuine concerns among researchers that fasting blood sugar alone, widely used in most population base surveys, may not capture all cases of undiagnosed diabetes in Africans, especially in the early stages of the disease. The high prevalence of diabetes mellitus among females in this study is consistent with the findings from a meta-analysis on the prevalence of diabetes mellitus in SSA, which reported a higher prevalence of diabetes among women in Southern African countries than men from the same region but the lower prevalence among women from Eastern, Central Africa, and West Africa countries than men of corresponding settings [38]. The high prevalence of DM among females in the SSA population is associated with the high prevalence of overweight/obesity.

The rate of smoking varies significantly throughout the world, with the prevalence rate of the female-to-male ratio directly related to the economic development of the country [39]. The women-to-men smoking prevalence ratio in high-income countries is ≈ 0.8, while in low and-middle income countries, the ratio is <0.1 [36,39]. Smoking rates differ considerably worldwide, and the ratio of women-to-men smoking prevalence rates is strongly related to a country’s level of economic development.[40]. In Africa, the prevalence of smoking is on the increase [36]. Despite the low smoking rate in this study, a surprisingly higher prevalence rate of women smokers (9.2%) than their male counterparts (7.5%) was noted, with the women-to-men ratio documented as ≈ 0.8. Our study is similar to reports from the Pacific Island state of Nauru, Denmark and Sweden [41]. While there is variance in smoking prevalence across Africa, the gender disparity of smoking documented in our study is an important risk factor for cardiovascular diseases. It, therefore, warrants more national policies on prevention and cessation programs in Sierra Leone.

Our study documented a high prevalence of physical inactivity among the study participants, with a significant sex-gender disparity of high physical inactivity among females than males. Consistent with existing literature, physical inactivity is relatively uncommon among African females [12,42]. Socio-cultural factors in SSA limit women’s outdoor activities, which explains the high prevalence of inactivity among females. Limited physical activities are a cardiovascular risk factor associated with deleterious health outcomes.

In the logistic regression analysis, we detected higher odds of dyslipidemia and alcohol consumption among women than men. In contrast, the multivariate analysis observed adjusted odds for hypertension, dyslipidemia, and alcohol for women than men. The odds of gender-specific cardiovascular risks were further evaluated in this study. Our findings observed a stronger correlation between BMI, WC, and raised blood sugar among women than men. In our study, we found gender differences to be consistent with other reported studies despite socio-cultural differences. [2,11,26,32,43].

This study underscores the burden of cardiovascular risk factor (CVRF) among adults and its sex-and-gender disparity for females in Sierra Leone. It further demystifies the misconception that females are less predisposed to cardiovascular diseases. Our findings have significant policy implications in guiding health promotion initiatives and lifestyle interventions, as female-specific cardiovascular risk factors may influence the impact and outcome of cardiovascular diseases.

## 5. Strengths and limitations

Our study has several strengths. First, the sample size is large, and the findings are representative. Second, we used accurate laboratory parameters and investigative tools (HBA1c), and trained clinicians collected data. Third, we used a structured data collection proforma (WHO Stepwise tool) that was pretested and validated before use. Finally, we adhered to ‘STROBE’ (Strengthening the Reporting of Observational Studies in Epidemiology) guidelines for reporting the study findings. The study’s limitations are as follows: Firstly, inference cannot be drawn from the causal relationships of the variable as the study is cross-sectional in design. Secondly, the generalizability of the study findings to Sierra Leone and other urban African settings is limited, as it represented adults residing in an urban setting. Despite these limitations, our study highlights important information regarding the prevalence and correlates of gender-specific CVD risk factors among adults in an urban African setting.

## 6. Implications and conclusions

The sex-and-gender disparity in cardiovascular diseases is of concern in Africa for the prevention, diagnosis, treatment and treatment of CVD. This study gives convincing evidence of the high prevalence of cardiovascular risks in Sierra Leone, especially among women, as the rate of obesity, diabetes and hypertension are rising steadily. The sex-and-gender disparity is mainly due to innate genes and environmental influences. Therefore, public health screening strategies, health education and promotion related to gender differences in cardiovascular diseases will help better prevent cardiovascular events, which may reduce the burden of CVD in Sierra Leone. Moreover, gender-related variables remain to be defined and added to current risk assessment in future studies.

## Data Availability

All data produced in the present study are available upon reasonable request to the authors

https://www.researchregistry.com/browse-the-registry#home/

## Data sharing statement

Data-set and statistical code can be shared on reasonable request to the corresponding author @jamesbwrussell@gmail.com

## ACKNOWLEDGMENTS

**The ECOBANK (SL) Study group:** Dr Jattu Rahman-Sesay, Dr. Tejan Mansaray, Dr. Jajuah, Dr. Mac-Jajuah, Dr. Mohamed Samura, Dr Abdul Karim Bah, Dr Evelyn Hawa Kamara, Dr Chernor Abubakarr Barrie, Paul Thoronka, Dr. Osman Kanneh, Dr. Alieu Kanu, Dr. Vidal Dupigny, Dr. Scholastica Nduisi, Dr. Omar Bah.

We thank all Ministry of Health and Sanitation nurses for serving as data collectors: Zainab Kargbo, Fatmata Koroma, Abigail Pratt, Claudia Campbell, Angel Jones, Lovetta Davies, Fatmata Bangura, Albert Rogers, Abibatu Jones, Alie Momoh and Gillian Jones. We are also grateful for the technical laboratory assistance provided by the Ecomed Advance Medical Laboratory and the Prime Care Medical Clinic staff, who supported the cardiac screening of the participants. We wish to thank all the participants enrolled in this study.

## Author’s Contributions

JBW Russell conceptualized, designed the overall study and formal analysis, and wrote – the original draft manuscript. S Sesay conceptualized and designed the overall research, TR Koroma, data curation, formal analysis writing and writing – original draft manuscript. SK Samura, data curation, and formal statistical analysis. A Bockarie, I.F. Kamara, LA Tetteh, Sorie Conteh, S Lakoh, OT Abiri, J.M Coker, DR Lisk, reviewing and editing. V Conteh, M Smith, project administration and community recruitment of participants. All authors were responsible for a critical review of the manuscript, for important intellectual content, and provided approval of the final draft.

## Informed Consent

The IRB cleared the informed consent, and every participant signed the informed consent before recruitment.

## Abbreviations

BMI: Body Mass Index
CMD: Cardiometabolic Disease
CMRF: Cardiometabolic risk factors
CVD: cardiovascular disease
CVDRF: cardiovascular disease risk factors
DBP: diastolic blood pressure
LDL-C: Low-density lipoprotein cholesterol
LMIC: low-and middle-income countries
LVH: left ventricular hypertrophy
LVMI: Left ventricular mass index
NCD: Non-communicable disease
HDL-C: High-density lipoprotein cholesterol
SSA: Sub-Sahara African
SBP: systolic blood pressure
SLE: Leones currency
TG: triglyceride
TOD: target organ damage
WC: waist circumference
WHO: World Health Organization

